# Natural history of retinitis pigmentosa based on genotype, vitamin A/E supplementation, and an electroretinogram biomarker

**DOI:** 10.1101/2022.11.26.22282587

**Authors:** Jason Comander, Carol Weigel DiFranco, Kit Sanderson, Emily Place, Matthew Maher, Erin Zampaglione, Yan Zhao, Rachel Huckfeldt, Kinga Bujakowska, Eric Pierce

**Affiliations:** Mass. Eye and Ear, Ocular Genomics Institute, Berman-Gund Laboratory for the Study of Retinal Degenerations, Harvard Medical School, Boston, Massachusetts

## Abstract

A randomized clinical trial that began in 1984 was conducted to determine the efficacy of vitamin A and E supplementation to reduce the rate of disease progression in patients with retinitis pigmentosa (RP). Vitamin A was shown to provide benefit while vitamin E had an adverse effect. Although genetic testing was unavailable at that time, banked DNA samples now provide the opportunity to combine modern genetic classifications with this extensively phenotyped longitudinal cohort. We hypothesized that the beneficial effects of vitamin A would vary by genetic subtype, and that the electroretinogram (ERG) 30Hz cone flicker implicit time could serve as a biomarker to predict disease progression. Existing genetic solutions or usable DNA samples were available for 96% of subjects. The overall genetic solution rate was 587/765 (77%) of sequenced samples. Combining genetic solutions with ERG outcomes produced a coherent dataset describing the natural history of RP among patients with multiple genetic causes of disease. There were systematic differences in severity and progression seen among different genetic subtypes of RP, confirming and extending findings made for disease caused by mutations in the most common causative genes, including *USH2A, RHO, RPGR, PRPF31*, and *EYS*. Baseline 30Hz flicker implicit time was a strong predictor of progression rate. Analyses using additional data from the original trial in combination with using the implicit time as a predictive biomarker showed the deleterious effect of vitamin E on progression was still present, but surprisingly found that the effect of vitamin A progression in the cohort as a whole was not detectable. Adding additional subjects from later trials to increase power gave similar results. Subgroup analyses among the largest gene groups revealed a potential adverse effect of vitamin A supplementation in patients with disease due to mutations in the *USH2A* gene and a trend toward benefit in patients with the p.Pro23His mutation in the *RHO* gene, based only on small groups. This study also demonstrated how genetic subtype and implicit time have significant predictive power for a patient’s rate of progression, which is useful prognostically. Validation of implicit time as a biomarker of disease progression, as demonstrated in this large cohort, may help with subject selection and endpoint selection in clinical trials for future experimental therapies. While vitamin E supplementation should still be avoided, these data do not support a generalized neuroprotective effect of vitamin A for all types of RP.

## Introduction

Retinitis pigmentosa (RP) is a slowly progressive, inherited, rod-cone retinal degeneration in which early rod photoreceptor death is typically followed by cone photoreceptor death, causing significant visual disability in most patients (1). Multiple strategies have been tested for treating RP spanning many decades(2-4). Some studies focused on the potential role of certain nutritional supplements in slowing disease progression (5-8), including three clinical trials that were conducted at our institution from 1984-2008 (9-11). The investigators from those three trials not only maintained and preserved complete databases of clinical trial data from all subjects but also created a comprehensive biobank for successful long-term storage of subject DNA samples. Now, 30 years later, this unique combination of clinical and genetic resources provides a rare opportunity to apply modern genetic and analytical techniques to a large cohort of RP subjects followed longitudinally.

The concept of the first trial (1984-1991), testing vitamin A and vitamin E supplementation to slow the progression of RP, was initially inspired by positive reports from patients who independently started taking one or both supplements (12). Formal dietary intake studies, combined with 3-year clinical data about progression rates, supported the hypothesis that vitamin A and/or E could be protective against the progression of RP (12). The roles of vitamin A and vitamin E in maintaining photoreceptor function were appreciated at the time as well. A 2×2 factorial design (“trace”, “A”, “E”, “A plus E”) was selected to efficiently test the role of two supplements, and their combination, in the setting of a masked, randomized clinical trial. The dose of vitamin A used was 15,000 IU of vitamin A palmitate per day. This trial was notably large (N=601 subjects) and lengthy (4-6 years of follow-up) for a rare disease (9). The primary outcome was the 30Hz cone flicker electroretinogram (ERG) amplitude, measured using signal averaging, bandpass filtering, and artifact rejection, to allow recording of smaller response amplitudes (9, 13). This cone flicker response amplitude shows a remarkably orderly exponential decay over a certain range of disease severity and correlates with clinically relevant outcomes, such as the ability to drive during the day or at night, walk alone at night, or be employed(14, 15).

The original results of the trial, that vitamin A supplementation slowed the progression of RP by 1.7% per year, and that vitamin E caused faster progression by 1.8% percent per year, were not met with uniform agreement (16-20). One criticism was that the largest effects of A and E were mostly seen in the last two years of the study (see Figure 5 of the original study), where the sample size was smaller (9). Regardless of this complexity, the addition of comprehensive genotyping to this dataset provided a relatively clean opportunity to test the hypothesis that vitamin A effect could differ depending upon the genetic cause of RP. One hypothesis is that some genetic subtype(s) benefited greatly, while others did not, resulting in the observed results seen in the ungenotyped RP cohort. For example, it could have been the case that RP associated with certain mutations in the rhodopsin gene would benefit from vitamin A treatment, while other genetic types do not. This hypothesis was motivated by the biochemical observations that vitamin A, which is a covalently bound cofactor of Rhodopsin, helps certain class II rhodopsin mutants to fold (21), and that vitamin A supplementation in mice expressing p.Thr17Met but not p.Pro347Ser mutant rhodopsin slowed progression of disease (22). In a different study, vitamin A supplementation was found to be adverse in a rhodopsin p.Asp190Asn mutant mouse line. Therefore, in this study, we tested the hypothesis that different genetic causes of RP might influence the responses to vitamin A and E supplementation. In the process of exploring this hypothesis, we had the opportunity to 1) determine genetic causes of disease in this well-characterized cohort, learning about the natural history of genetic subtypes of RP, and 2) re-examine details of the results of the original vitamin A/E study.

The original investigators did look for potential differential treatment responses among the “genetic subtypes”, which at the time were defined as recessive, dominant, X-linked, or unknown, based on pedigree analysis and limited molecular genetic solutions. No differential treatment responses were identified using these categories (9). With the intervening revolution in genetic sequencing, this study applied modern targeted DNA resequencing of all known IRD genes combined with expert variant annotation to reveal genetic diagnoses for a large fraction of previously-unsolved subjects in the study (23, 24). We also incorporated a recently defined biomarker for the progression of 30Hz cone flicker ERG amplitudes, namely the baseline cone flicker implicit time, as a predictive variable (25, 26). Because the results from re-analyzing the original vitamin A/E trial were different than expected (see Results), additional subjects were added for increased statistical power from the vitamin A-only arms of two later clinical trials that tested the effect of DHA supplementation or lutein supplementation (10, 11). Using this approach, we re-evaluated the conclusions of the original study and produced a coherent dataset describing the natural history of RP among different molecularly defined genotypes.

## Methods

### Cohorts

From 1984 to 1991, 601 patients participated in the vitamin A/E trial (NCT00000114) (12). Patients with certain forms of “atypical RP” were excluded, such as Usher syndrome type I, Bardet-Biedl syndrome, pericentral RP, sector RP, and X-linked RP carriers. Usher syndrome type 2 patients were included, and syndromic versus non-syndromic presentations were not distinguished for purposes of analysis. Patients in all 4 treatment groups (“trace”, “A”, “E”, “A plus E”) were included in the current analyses. Preliminary analyses of the effect of vitamin A and E on longitudinal progression rates were different than the findings of the original study (see Results), and therefore additional subjects were added from later clinical trials to increase the sample size. Specifically, subjects were added from the vitamin A-only controls arms of the subsequent, separate DHA trial (N=91, NCT00000116) and lutein trial (N=107, NCT00346333) (10, 11). The subjects who received DHA or lutein were not included, neither in the DNA resequencing efforts nor in the data analysis.

We assessed for homogeneity of the three data sets (See supplemental Methods). This model showed that, compared to the vitamin A trial, subjects in the lutein trial had a similar progression rate (beta = 0.01, p=0.59), but subjects in the DHA trial showed a faster progression rate (beta = -0.02, p=0.039). Therefore the subjects from the DHA trial were not used for the purposes of regression modeling of rates of decline. However, the data from all three trials were used elsewhere in this study, where the rate of decline is not explicitly being modelled.

### Clinical Data

The methods for data collection for all three trials can be found in the original publications (9-11). We note that the ERG data acquisition methods and equipment were carefully maintained over the years to ensure consistent measurements (and remain in clinical use). These methods were developed before ISCEV protocols and use a Xenon flash of 0.2 cd*s/m^2^ instead of the now-standard 3 cd*s/m^2^ flashes for recording the 30Hz flicker responses. The recording acquisition and processing also use signal averaging, bandpass filtering, and artifact rejection, to allow recording of smaller response amplitudes (13). Goldmann central visual field areas were measured by planimetry of the V4e response area that was contiguous with the center, and converted to an equivalent circular diameter using the formula diameter = 2 * sqrt(area/pi).

### Genotyping

Pre-existing genetic solutions were available for 211 samples. 554 unsolved cases, for whom DNA samples were available, were analyzed with the Genetic Eye Disease (GEDi) targeted sequencing panel of all known IRD genes, as described previously (23), or by Sanger sequencing in 4 cases. Sequence data was aligned to the hg38 genome build and the subsequent variant calling, annotation, and analyses were performed as described (24). Copy number variation (CNV) predictions were produced using gCNV (27, 28), and the known *MAK*-Alu structural variant was identified using a custom script (29). Screening of mutations in the *RPGR* ORF15 was performed by PCR amplification of the target region and Sanger sequencing using established methods (30). Variants were classified according to the American College of Medical Genetics and Genomics (ACMG) guidelines (31, 32) and adjudicated by the co-authors.

Recessive solutions were required to have two mutations identified, but segregation testing was not available in many cases. Samples were marked as either solved or unsolved, with the solved samples being pooled by gene into gene-specific groups for further analysis. The small number of samples without sequencing data available (N=34) were also marked as unsolved. Some solutions have been previously published (see Supplementary Table 1). 61 of the 765 solutions (8%) were obtained from whole exome sequencing as published (24, 28).

### Longitudinal Analysis of ERG data

Data processing of ERG parameters was performed as closely as possible to the original study in consultation with the original data manager of the study (CW) (9). For additional description of ERG data processing and statistical methods, please see Supplemental Methods.

Over the course of several iterations of data processing (e.g. as more subjects were solved genetically and added), it was noted that when looking at small subsets of subjects, there were unstable estimates of the effects of vitamin A and E on progression rates with small changes in the input dataset (not shown). Power calculations further supported that very small subgroups should not be analyzed.

Therefore analyses for single gene subgroups were only performed for the larger gene groups: *USH2A, RHO, RPGR, PRPF31*, and *EYS*. Genetic solutions for smaller gene groups were pooled into an “Other solutions” group for the regression models. The “Unsolved” group represents the group of subjects who had no genetic solution identified after sequencing and analysis. In general, all mutations in the same gene were pooled together; however, an additional hypothesis was made that certain mutations or mutation classes of *RHO* mutations might be differentially responsive to vitamin A (22). There was only one subgroup, RHO P23H, with enough patients (N=19) to attempt a subgroup analysis.

There are some subjects with additional long-term follow-up data that is now available, including those who participated in multiple trials and accumulated very long total follow-up time. Initial regression modeling using imbalanced very long-term data (e.g. 23 years) from some subjects and only 4-6 year data from the rest of the subjects produced results that were overly weighted by the outcomes of the very long-term subjects (not shown). It also would have been complex to model subjects who switched vitamin A treatment status during these longer follow-up time periods. Therefore, for each subject, only the first 6 years of data were used for regression models.

Subsequent analyses were performed in R; see Supplemental Methods.

### Study Approval

The research was conducted in compliance with the Mass Eye and Ear Institutional Review Board (IRB) approval. Written informed consent had been obtained from the subjects after explanation of the nature and possible consequences of the study.

## Results

### Genetic solutions

Pre-existing genetic solutions from prior studies were known for 211 of the 799 subjects included in these analyses. Of the remaining 588 subjects, sequencing of banked DNA samples, which generally were in storage for 15-30 years, was highly successful. Of the 588 subjects without existing solutions, 554/588 (94%) had usable DNA samples available, which were used for next-generation sequencing (N=550) or targeted Sanger sequencing when a familial solution was known (N=4). Including the pre-existing solutions and including subjects who could not be sequenced, the solution rate was 587/799 (73%). Excluding subjects with no remaining usable DNA sample, genetic solutions for 587/765 (77%) sequenced subjects were found. The genetic solutions spanned 53 different genes (Figure 1, Supplemental Table 1). The genetic solution rate (solved samples / sequenced samples) was similar between the three different study sources analyzed: Vitamin A 77%, DHA 72%, Lutein 77%. The most common causative gene was *USH2A*, found in 136/587 (23%) of the subjects with solutions. There were 19 genes identified as the cause of disease in only a single subject.

**Figure 1.**
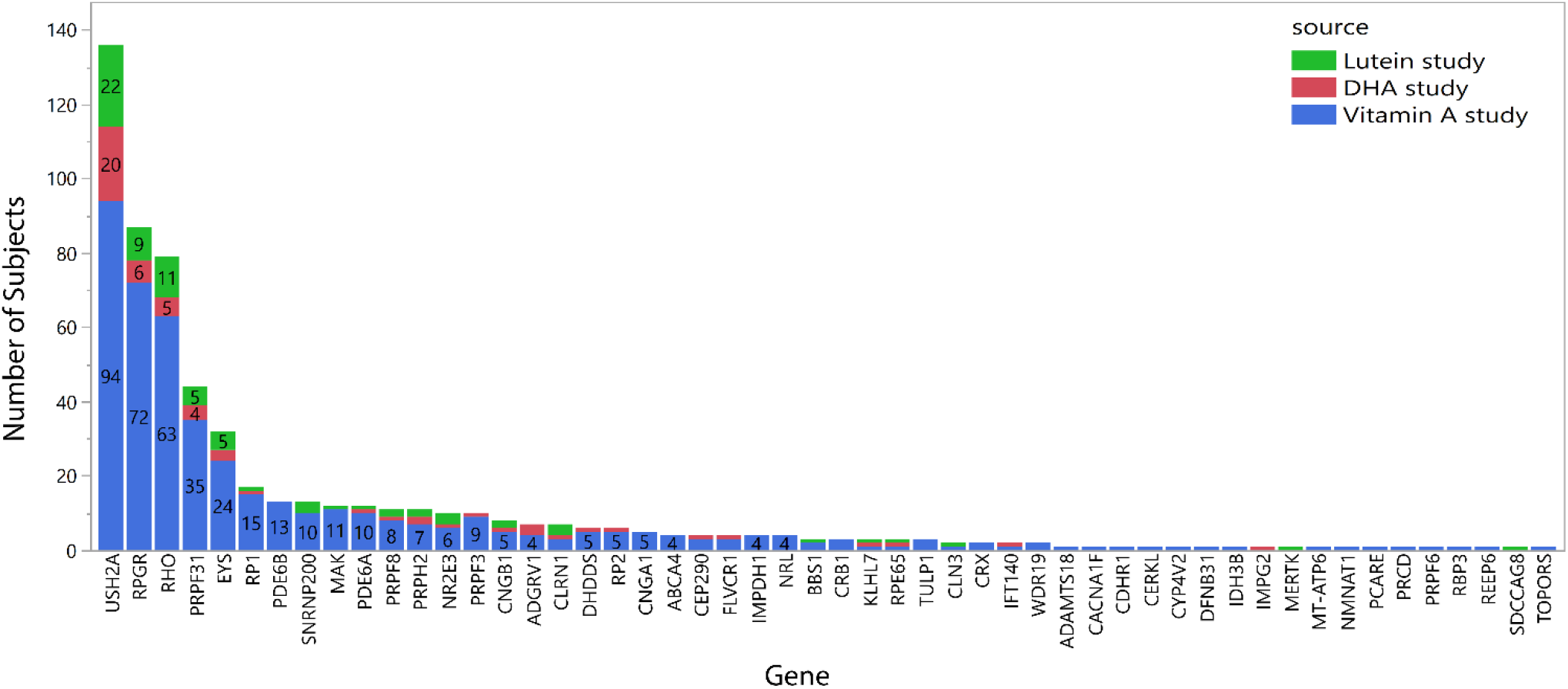
A histogram shows the number of subjects with a genetic solution in each gene. Bars are labeled with the number of subjects. Subject counts are reported separately for each study because the subjects from the vitamin A study were recruited before any genetic solutions for RP were known, and therefore the distribution of genes in that study should be the most unbiased.

### Natural History

Baseline characteristics were analyzed by gene for subjects with mutations in gene groups with ten or more subjects (N=487 subjects, 14 genes; Table 1). Data from less common gene groups (N<10 subjects) or unsolved samples are not shown in Table 1. Subjects with *RPGR* mutations had their first clinical visit at the youngest age, reflecting a more severe phenotype, while subjects with mutations in *MAK* were oldest, mirroring their good visual acuity and visual fields. Subjects with *PRPH2* and *NR2E3* mutations had the best visual fields, whereas subjects with mutations in *PRPF3, PDE6B*, and *RPGR* had the worst visual fields. Subjects with *RPGR* mutations also had the lowest visual acuity. While a higher ERG amplitude is generally correlated with a shorter implicit time, subjects with mutations in *EYS* had particularly short implicit times, paired with some of the lowest 30 Hz ERG amplitudes. Conversely, *SNRNP200* subjects had long implicit times with relatively high 30 Hz ERG amplitudes, based on N=13 subjects only. Although the severity of visual field and 30 Hz ERG amplitudes showed correspondence for many groups, there were also examples of one of these measures being disproportionately large relative to the other, for example in *PRPF31*. (These trends are comparisons between the gene group mean values and are not intended to represent all individual patient values which are spread within each gene group.)

**Table 1.** Baseline characteristics are shown for the largest genetic subgroups. Values represent: Average ± standard deviation (N). Color coding of each column highlights the relatively more or less severe phenotypes among gene groups.

| Gene group | Age | Visual acuity<br>(dec) | Central GVF<br>equiv.<br>diameter<br>(deg., V4e) | ERG cone<br>flicker<br>amplitude<br>(microvolts) | ERG cone<br>flicker implicit<br>time<br>(microsec) |
| --- | --- | --- | --- | --- | --- |
| USH2A | 36 ± 8 (136) | 0.72 ± 0.21 (135) | 82 ± 52 (95) | 3.9 ± 6.4 (134) | 43.1 ± 3.9 (136) |
| RPGR | 28 ± 7 (87) | 0.47 ± 0.23 (87) | 52 ± 39 (73) | 1.8 ± 1.9 (78) | 44.5 ± 3.1 (87) |
| RHO | 35 ± 8 (79) | 0.74 ± 0.23 (79) | 72 ± 54 (64) | 9.9 ± 14 (79) | 40.9 ± 4.5 (79) |
| PRPF31 | 33 ± 8 (44) | 0.68 ± 0.26 (44) | 82 ± 54 (35) | 3.6 ± 4.6 (44) | 43.7 ± 4.2 (44) |
| EYS | 36 ± 8 (32) | 0.72 ± 0.18 (32) | 85 ± 46 (24) | 2.6 ± 5.3 (32) | 41.9 ± 4.3 (32) |
| RP1 | 38 ± 8 (17) | 0.82 ± 0.15 (17) | 72 ± 58 (15) | 5.1 ± 6.9 (17) | 41.3 ± 3.5 (17) |
| SNRNP200 | 33 ± 8 (13) | 0.72 ± 0.21 (13) | 87 ± 62 (10) | 4.7 ± 11 (13) | 44.2 ± 3.7 (13) |
| PDE6B | 35 ± 6 (13) | 0.66 ± 0.19 (13) | 48 ± 33 (13) | 6.2 ± 11 (11) | 42.8 ± 5.1 (13) |
| PDE6A | 30 ± 8 (12) | 0.70 ± 0.29 (12) | 85 ± 61 (9) | 7.6 ± 11 (12) | 43.1 ± 3.9 (12) |
| MAK | 39 ± 7 (12) | 0.82 ± 0.17 (12) | 85 ± 54 (11) | 3.5 ± 4.3 (12) | 42.9 ± 2.3 (12) |
| PRPF8 | 31 ± 8 (11) | 0.70 ± 0.29 (11) | 59 ± 39 (8) | 1.8 ± 2 (10) | 43.3 ± 4.6 (11) |
| PRPH2 | 37 ± 10 (11) | 0.80 ± 0.23 (11) | 94 ± 64 (7) | 6.3 ± 7.3 (11) | 42.7 ± 3.0 (11) |
| PRPF3 | 34 ± 9 (10) | 0.57 ± 0.23 (10) | 47 ± 33 (10) | 1.0 ± 0.75 (10) | 43.2 ± 5.8 (10) |
| NR2E3 | 35 ± 9 (10) | 0.63 ± 0.18 (10) | 90 ± 41 (7) | 3.6 ± 5.1 (10) | 43.5 ± 4.4 (10) |

Figure 2 shows how visual acuity, central visual field diameter, and ERG cone flicker amplitude are related to age, in the five largest genotype groups. These graphs demonstrate the heterogeneity between different subjects even within gene groups. In general, all groups tend to lose visual field at a younger age, and acuity at a later age, especially the *USH2A* group; this is reflected in the cluster of lines appearing further to the left for the visual field row, and further to the right/center in the acuity row.

**Figure 2.**
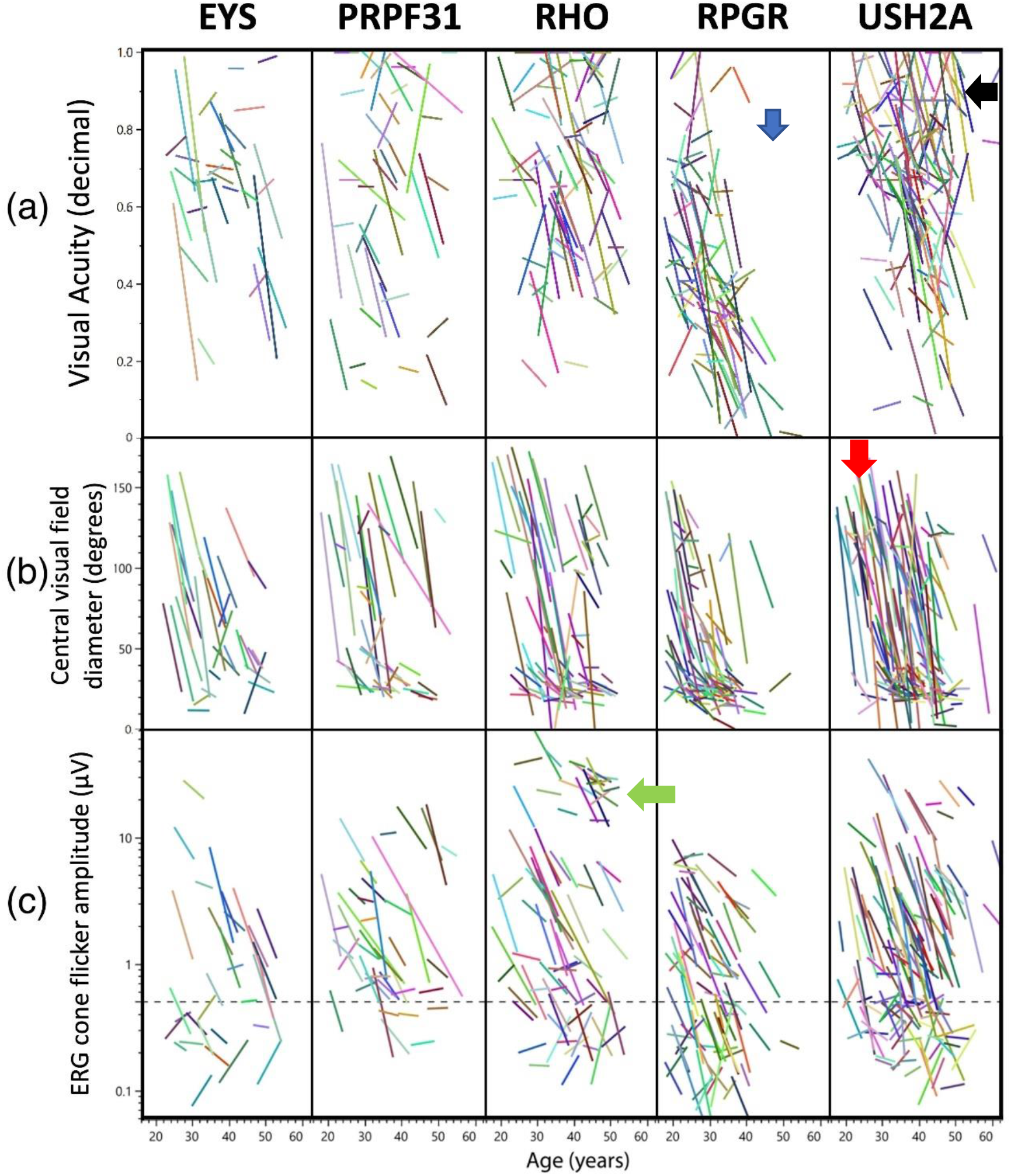
Visual acuity (a), central visual field equivalent diameter (b), and ERG cone flicker amplitude (c) are shown for the five largest genetic subgroups. A linear curve fit is shown for the data for each subject. The dashed line in C represents 0.5 microvolts, below which the decay of the response amplitude is less reliably estimated. The lack of points below the blue arrow demonstrates the absence of *RPGR* subjects with normal visual acuity after age 40. *USH2A* subjects have relatively steep visual field declines starting at a variety of ages (red arrow) but can maintain visual acuity into older ages (black arrow). A subset of *RHO* subjects has particularly mild deficits in the electroretinogram amplitudes even at older ages (green arrow).

Further gene-specific observations are described in Figure 2.

### Effects of vitamin A/E supplementation on ERG cone flicker progression rates

Modeling of the effects of vitamin A and vitamin E on the rate of ERG progression using the subjects from the original vitamin A/E study initially produced different results than the original publication; vitamin A had a beneficial significant effect (Table 2, “base model”, p=0.004), with a slightly smaller p-value than that seen in the original paper (p<0.001). Vitamin E, which had a significant negative influence in the original study (p=0.04), also showed a negative trend in Table 2 (“base model”, p=0.06). Reconciliation of available datasets revealed that the data in the “base model” includes additional year 5 and 6 data that was obtained after the data lock used in the original study, which excluded data mostly after September 1991. (Furthermore, a slightly different subset of subjects met the minimum baseline ERG amplitude requirement when some minor data processing variabilities in the original data were strictly standardized, e.g. averaging OD and OS values before applying the minimum amplitude cutoff.) Using the additional post data lock data (i.e. all available year 0-6 data) lowered the size of the vitamin A effect and removed the statistical significance of the vitamin E effect. Because the data at years 5 and 6 still had smaller numbers of subjects than in prior years, we repeated the analysis on years 0-4 only using model #1. Vitamin A had no significant effect (p>0.05) and vitamin E had a borderline negative effect (p=0.046) when looking at the 0-4 year data. Further analyses were performed with all available year 0-6 data (Table 2).

**Table 2.**
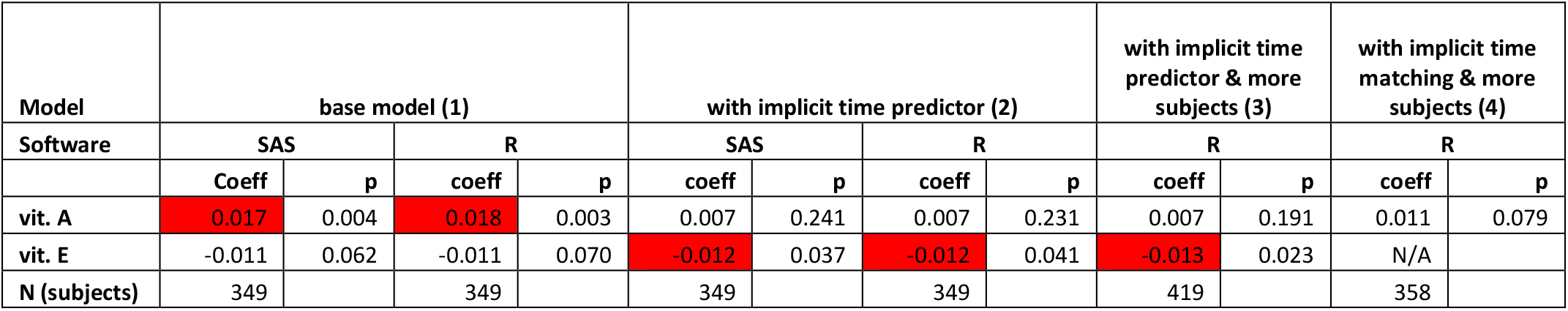
Vitamin A and E treatment effects in the original vitamin A trial cohort, modeled over years 0-6. A mixed model was used to model the yearly rate of decline (beta coefficient, “coeff”) of the loge ERG amplitude (model 1). A coefficient of -0.01 represents approximately a 1% decline per year of remaining function. The “with implicit time predictor” model #2 adds baseline implicit time as a predictor of ERG decline. With the additional predictor and with the post-data lock data, the observed vitamin A effect was diminished further and is no longer significant; however, the vitamin E effect persists. Model #3 uses the same variables as model #2 but adds additional subjects from the lutein clinical trial. Model #4 uses propensity matching to select a subset of data with balanced baseline implicit times between A+ and A-treatment groups, with similar results.

Next the baseline implicit time was added to the regression model as a predictor of rate of progression (Table 2, model #2 “with implicit time predictor”). This biomarker was highly predictive, showing an additional ERG amplitude decline of -0.01 log_e_ units (∼1%) per millisecond of baseline implicit time compared to an overall -0.1 log_e_ unit (∼10%) yearly decline in the model without implicit time. A previous study showed implicit time was predictive for *RHO* RP patients (26). In this dataset, the baseline implicit time was significantly predictive of rate of decline in general, and also within *RHO, RPGR*, and *USH2A* subgroups when analyzed separately (p<0.05 for each). The relationship between ERG decline rate and baseline implicit time is shown graphically in Figure 3. Using a mixed model with implicit time as a predictor, there was no significant effect of vitamin A (p>0.05) and a negative effect of vitamin E of -0.012 log_e_ units (∼1.2%), p=0.04. Part of the vitamin A effect had already been lost by using additional post data lock data from years 5-6 (see Table 2 model #1). When also using implicit time as a predictor (model #2), the remaining vitamin A effect was lost because, by coincidental imbalance at randomization, the vitamin A+ and A-arms were imbalanced for implicit time at baseline, with median baseline implicit times of: 42.8 msec for A+/E± groups and 44.2 msec for A-/E± groups (p=0.007). (The influence of implicit time as a predictive biomarker was not known at the time of the trial.)

**Figure 3.**
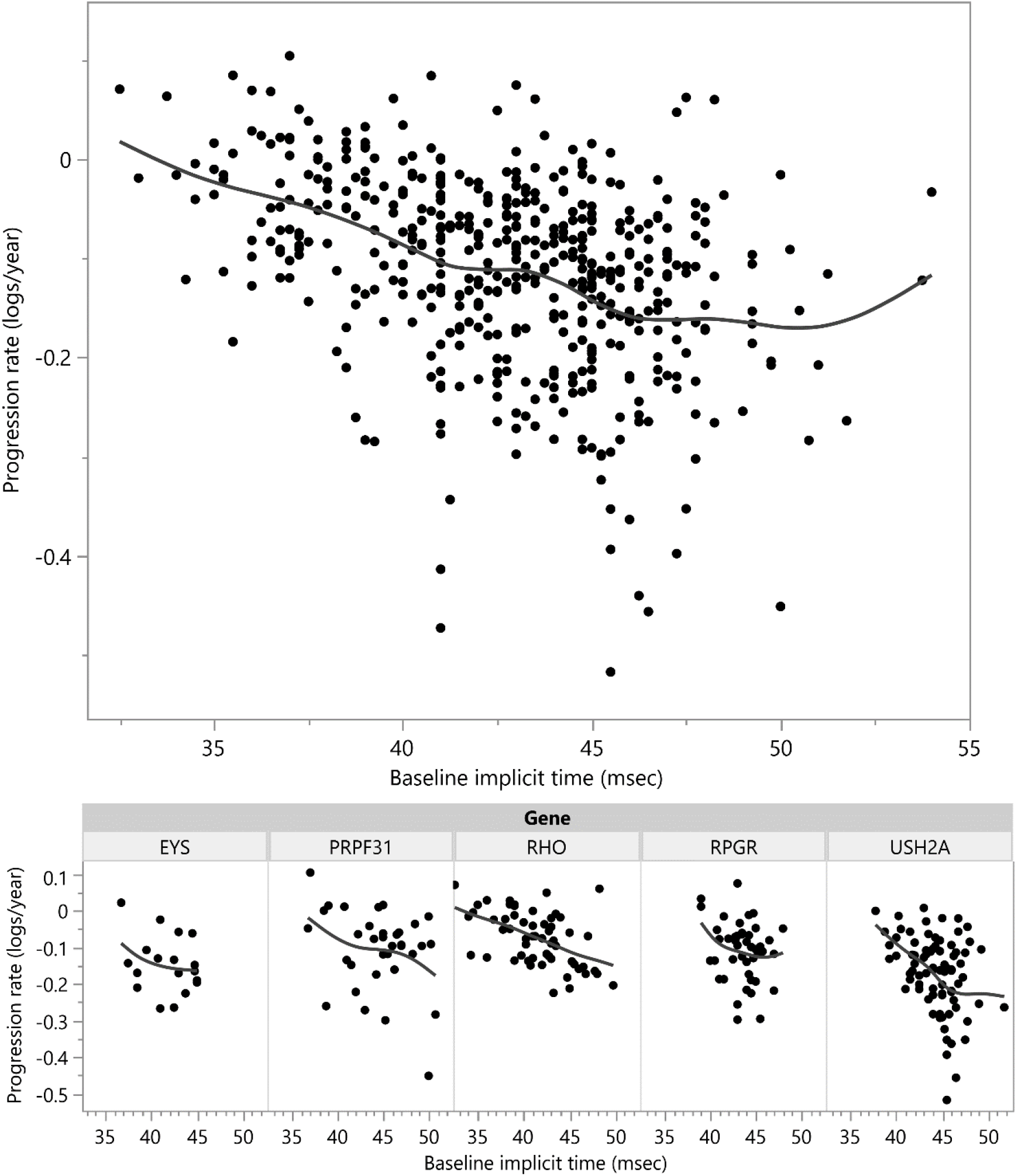
Individual cone flicker ERG amplitude decay rates were calculated for each subject (y-axis), and plotted by baseline ERG cone flicker implicit time (x-axis), for all subjects (top). A spline fit shows the trend towards worse progression rates with increasing baseline implicit time. Subjects from the largest gene subgroups are shown below. (One outlier point beyond the y-axis is not shown.)

Having noted that the original conclusions were not reproduced with these new analyses, we expanded the sample size by adding additional subjects taking vitamin A only from the two *control* arms of the subsequent DHA and lutein trials (10, 11). The subjects in the *active* arms of those trials, who took either DHA or lutein, were not included for sequencing or analysis, to avoid adding additional treatment variables. The total dataset consisted of 799 subjects from all three trials. It was only possible to add subjects to the A+ / E-arm of the analysis, as subjects in the subsequent trials had been advised to take vitamin A and avoid vitamin E supplementation based on the results of the original trial. Due to this imbalance, the analyses are presented both without (model 2) and with (model 3) these extra subjects. The subjects from the DHA trial showed a faster rate of decline (see Methods) and therefore were not used in this rate-of-change analysis (although including or excluding those subjects made only trivial differences, not shown). After adding additional subjects from the lutein trial (Table 2 model #3 “with implicit time predictor and more subjects”), there was no significant effect of vitamin A (p>0.05), but the adverse effect of vitamin E persisted (−0.013 log_e_ units, ∼1.3%/year, p=0.02). Interaction terms between vitamins A and E were not statistically significant for any of the above models.

To assess if the choice of statistical packages and models contributed to these findings, the analyses were implemented in the R Lmer package (See Methods). Nearly identical results were obtained between SAS and R (Table 2). To assess for the possibility that there is hidden collinearity between baseline implicit time and assigned vitamin A treatment group, and to test for vitamin A effect in a smaller model with the minimal number of variables, propensity score matching was used to create a balanced dataset where baseline implicit times are matched between groups. In other words, a subset of the data was used to create a balanced case-control style analysis instead of using all subjects and numerically controlling for the imbalance in implicit times between the groups. The matched dataset (N=358 subjects, range 40-139 subjects per group) showed homogeneity of baseline implicit time between groups (p=0.97). With the balanced dataset, there was still no beneficial effect of vitamin A treatment (Table 2, model #4 “with implicit time matching & more subjects”).

In summary, adding additional year 5 and year 6 data, in combination with adding the predictive power of baseline implicit time to adjust for an imbalanced randomization at baseline, removed the evidence for the broad, beneficial effect of vitamin A that was seen in the original study. This lack of a significant vitamin A treatment effect persisted after adding additional subjects from a later trial. The negative effect of vitamin E became smaller but remained statistically significant.

### Rates of decline, by gene, and with vitamin A/E supplementation

Next, genotype information was added to the progression models. For this purpose, “genotype-specific” is defined at the gene level, pooling together all subjects with mutations in the same gene. Small gene groups were pooled into an “Other solutions” group to avoid spurious values from small subgroups (see Methods). A regression model was created estimating the cone flicker ERG amplitude and the cone ERG amplitude progression rate over time, with gene groups of *EYS*, Other, *PRPF31, RHO, RPGR*, Unsolved, and *USH2A*. The effect of the gene on amplitude and on the progression rate of amplitude was strong (p<0.00001, p<0.00001, respectively, type 3 test). Figure 4 and Supplemental Table 2 show the rates of progression across the five largest genotype groups. Among the subjects who started the study with sufficient ERG amplitude to measure change over time (N=419), subjects with mutations in *EYS* and *USH2A* showed the highest progression rates, while *RHO* was the slowest. This observation was consistent whether or not baseline implicit time was controlled for. In order to determine the comparative importance of the different variables in influencing progression rate, calculation of standardized beta regression coefficients showed that there were similar effect sizes for gene effects and for the baseline implicit time effect (not shown). This indicates that gene effects and implicit time effects are of similar importance in predicting progression rate.

**Figure 4.**
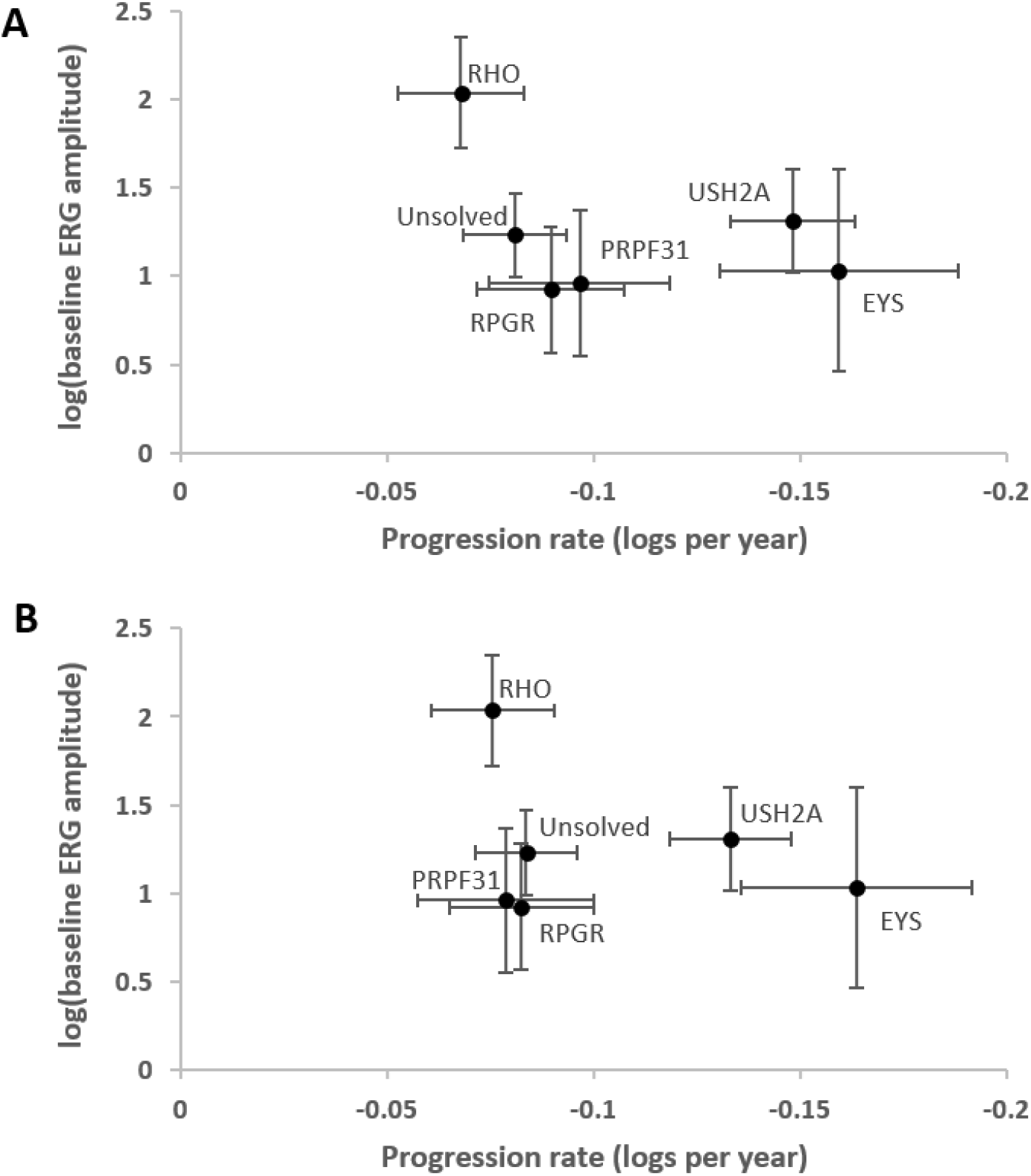
Gene-specific estimates of the baseline (time=0) cone flicker ERG amplitude (y-axis) and the yearly ERG amplitude progression rate (x-axis). Both x- and y-axis values are adjusted to remove any vitamin E treatment effect. The results are presented without (A) and with (B) adjustment for baseline implicit time on the x-axis. A negative value on the x-axis represents a decrease in amplitude over time.

Note that because of measurement variability at very low ERG amplitudes, there is a “floor effect” beneath which progression cannot be assessed accurately. Therefore these progression models only include subjects (N=419) whose baseline ERGs were high enough to accurately determine rates of decline (See Methods). This selection of subjects with a minimum starting amplitude causes all gene groups except *RHO* to artificially cluster on the y-axis “floor” at a starting log_e_(baseline ERG amplitude) of ∼1-1.5; therefore estimates of baseline disease severity should be obtained from the “Natural History” section above, in which there was no lower limit for inclusion in the analysis. The intrinsic progression rate, however, is shown on the x-axis and demonstrates the differences in progression between gene groups.

Table 3 shows the results of a regression model used to estimate the effects of vitamin A and vitamin E on progression rate within the genotype subgroups. When looking at the effect of treatment on progression rate, there was a significant interaction with the gene variable overall (p=0.002). This justifies looking at the gene subgroups. There were no subgroups in which vitamin A showed a beneficial effect (except the “Unsolved” group, which does not have a specific biological meaning). In the *USH2A* group (N=65 subjects), an adverse effect of vitamin A treatment was observed (coeff = -0.039, p=0.03). A borderline adverse effect (p=0.054) was seen in the *EYS* subgroup, based on a small sample size. If the same analysis is repeated with outlier removal for baseline implicit time, the coefficients and p values showed only minor changes, including a p-value of 0.047 instead of 0.054 for the vitamin A effect in the EYS subgroup. Because this finding is based on a very small sample size (N=15 subjects across all arms), the results from the *EYS* group should be viewed with additional caution.

**Table 3.** Vitamin A and E treatment effects modeled in subgroups based on genetic cause of disease. A mixed model was used to model the yearly rate of decline (coefficient, “coeff”) of the log_e_ ERG amplitude, in the vitamin A and lutein trial cohorts. Coefficients with significant p-values are highlighted in red. The USH2A subgroup shows a deleterious effect, and the Unsolved group shows a beneficial effect of vitamin A. A power calculation of >0.8 indicates sufficient power to detect an effect with the specified subgroup size. P values are corrected for multiple comparisons.

|  | EYS |  | PRPF31 |  | RHO |  | RPGR |  | Unsolved |  | USH2A |  |
| --- | --- | --- | --- | --- | --- | --- | --- | --- | --- | --- | --- | --- |
| Term | coeff | p | coeff | p | Coeff | p | coeff | p | coeff | p | coeff | p |
| vit. A | -0.066 | 0.054 | -0.010 | 0.675 | 0.019 | 0.315 | -0.015 | 0.456 | 0.031 | 0.032 | -0.039 | 0.032 |
| vit. E | -0.050 | 0.196 | -0.041 | 0.196 | -0.007 | 0.648 | 0.016 | 0.416 | -0.018 | 0.196 | -0.025 | 0.196 |
| N (subjects) | 15 |  | 31 |  | 52 |  | 39 |  | 102 |  | 65 |  |
| Power | 0.98 & 0.85 |  | 0.13 & 0.95 |  | 0.55 & 0.12 |  | 0.30 & 0.34 |  | 1 & 0.79 |  | 1 & 0.89 |  |

An additional hypothesis was made that certain *RHO* mutations might be particularly amenable to vitamin A chaperone therapy while others might not be responsive (22). Among the 52 *RHO* subjects the largest groups were p.Pro23His (N=19), p.Pro347Leu (N=4), p.Arg135Trp (N=3), while all other mutations had N=1 or 2. Therefore only the p.Pro23His group (henceforth, “RHO P23H”) was considered further.

A regression model was constructed with RHO P23H as a separate gene group (N=19), along with the other gene groups shown in Table 3 (N=419 total). The results in Table 4 show a trend towards a beneficial effect of vitamin A treatment in this subgroup (+0.063 log_e_ units, ∼6.3% per year, N=19 subjects), though with correction for multiple comparisons (N=8 groups), this effect is not statistically significant (p=0.063). Vitamin E was still adverse, though not significantly, in this model. While, the power calculations are relatively high for this subgroup and the magnitude of the trend is relatively large, similar to the EYS subgroup finding above, the RHO P23H finding should be viewed with additional caution in this small subgroup (N=19 subjects across all arms).

**Table 4.**
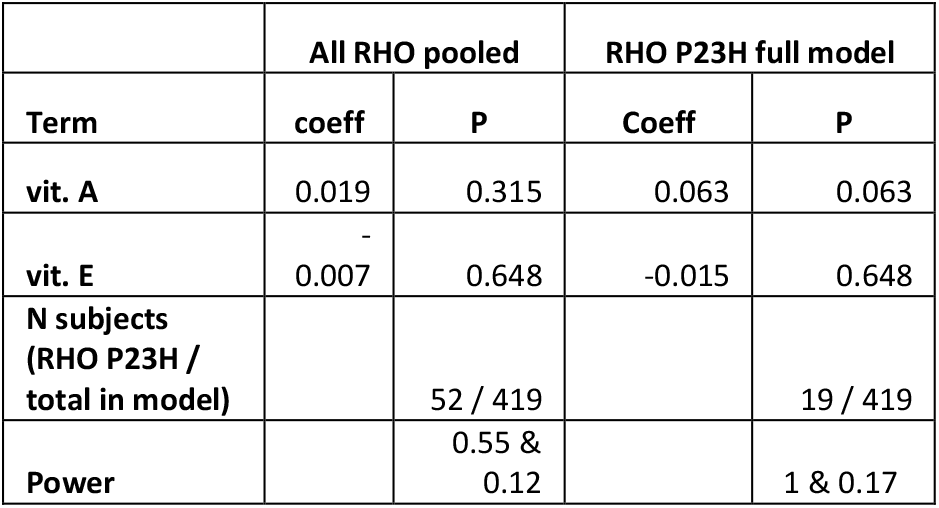
Vitamin A and E treatment effects modeled in the RHO P23H subgroup (right). A mixed model was used to model the yearly rate of decline (coefficient, “coeff”) of the log_e_ ERG amplitude, in the vitamin A and lutein trial cohorts. A power calculation of >0.8 indicates good power to detect an effect with the specified subgroup size. The RHO P23H subgroup showed a trend towards a beneficial effect with vitamin A treatment (p=0.063, right). For reference, all RHO subjects of any mutation pooled together showed no effect of vitamin A or E treatment, using the same analysis as Table 3 (left).

## Discussion

This study clearly demonstrates the variety of severities and progression rates that occur in RP, based on a well-documented cohort of patients with RP with longitudinal data. Having successfully determined a molecular cause of disease for 73% of all subjects (587/799) evaluated, we also demonstrate the success of what may be the oldest DNA biobanking effort for this rare disease.

Combining this high-quality phenotype and genotype data improves our understanding of the severity and progression of various RP genetic subtypes. This data can be useful both for prognostic information for patients, as well as for planning patient selection and endpoints for interventional clinical trials. We also found that in contrast to the original reports, updated analyses did not show a benefit for vitamin A supplementation in reducing progression of disease for patients with RP.

### Natural History of RP Among Major Genotypes

As enrollment in the vitamin A study was performed without knowledge of the underlying genotypes, it provides an unbiased view of the genetic composition and natural history of RP. Some genetic causes of RP had been discovered by the time the additional subjects were recruited for the lutein and DHA trials, but panel-based genetic testing had not yet been implemented. There were recruitment efforts for *RHO* families in the department during the time period of the vitamin A studies. Therefore, the distribution of genes in the DHA and lutein trials in Figure 1, which make up a small part of the overall dataset, may be slightly skewed by these efforts. For this reason the gene counts for each trial are reported separately. However this potential bias may be limited by the study design to include only one subject from each family (10, 11).

Starting amplitudes and rates of decline are shown in Figures 3 and 4. While the natural histories for the 4 largest groups (*USH2A, RPGR, RHO, PRPF31*) have been published using data from our institution (33-36), this dataset, which uses some of the same subjects, integrates the findings into a single model where cross-comparison between the groups and to the “average” case of RP is straightforward. We additionally provide comparable progression rates for RP caused by mutations in *EYS*, the next-largest group in this dataset. Baseline severity data for the 14 largest genotype groups is listed in Table 1.

Specifically, it is worth noting that rhodopsin-associated RP remains the mildest among the major genotypes, with a higher baseline ERG and a slower rate of progression. While there are differences in severity among different rhodopsin mutations (35), the average severity of the group as a whole is quite mild in comparison to other genotypes. The *RPGR* subgroup also continues to stand out as the most severe genotype among the large groups (Table 1), across nearly every metric. Mutations in genes that encode RNA splicing factors including *PRPF31, SNRNP200, PRPF8* and *PRPF3* are a common cause of IRD accounting for 76 subjects in total; PRPF3 and PRPF8 cause relatively severe disease (Table 1). *EYS* subjects had comparatively low ERG amplitudes despite having a faster (protective) cone flicker implicit time. *EYS* subjects also had a surprisingly fast average progression rate, which was the fastest among the top five gene subgroups. The progression rate showed a trend toward faster progression with vitamin A or E treatment, which complicates the estimation of the true rate without supplementation. It will be interesting to see whether this faster rate of decline is captured in the measures being assessed in the larger cohort of individuals studied in the ongoing natural history study of *EYS* subjects (clinicaltrials.gov NCT04127006), and in a similar prospective natural history study of *USH2A* (clinicaltrials.gov NCT03146078). While a faster progression rate is a poor prognostic sign for affected *EYS* patients, it may also provide an opportunity to plan shorter clinical trials for interventions which have the potential to slow progression of full-field metrics like the full field ERG.

One limitation of these progression models such as those in Figures 3 and 4 is that they can only be used with data from subjects with high enough baseline ERG amplitudes so that a decline can be reliably measured over time. In the *RPGR* group, nearly half (48%) of the subjects start with very low ERGs in which progression cannot be assessed. Therefore, the progression rates in the most severe groups are unavoidably based on unusually mild subjects in such groups. Of note, this issue does not apply to the cross-sectional data in Figure 2, which includes all subjects regardless of initial ERG amplitude. The clinical trials included only subjects with “typical RP”, and therefore the *RPGR* findings do not include subjects with cone-rod dystrophies or cone dystrophies.

It was also notable that subjects with *NR2E3* associated RP, comparatively, had particularly good visual fields. Indeed it is interesting that 10 *NR2E3* subjects were included in this cohort of “typical RP” patients, despite a history in our department of attempting to separate out the Enhanced S-cone Syndrome / Goldmann-Favre phenotype into a separate category based on factors such as the appearance of clumped pigment and blue-on-yellow visual field testing (37). Similarly, four ABCA4 subjects were included in the cohort. While ABCA4 defects can cause RP, they more typically cause an inherited macular degeneration (Stargardt disease) with varying degrees of full field cone-or cone-rod dystrophy. While vitamin A processing is known to be defective in this genotype (38), only one ABCA4 subject had sufficient starting ERG amplitude to measure a progression rate, so no comparisons of progression between vitamin A treatment groups is possible in *ABCA4* subjects. Further “unexpected” genetic findings in Supplemental Table 1, such as the presence of two manifesting *RPGR* female carriers, and “dominant” pedigrees that turned out to have X-linked disease, have been previously described in other cohorts (39, 40).

This study did not estimate comparable progression rates by genotype for other outcomes including visual field, visual acuity, or the mixed-response full field ERG; comparing group differences and progression rates in these outcomes could be an area for future research. Additional areas of study could be comparison of syndromic vs nonsyndromic disease(41-43) and expanding upon genotype/phenotype relationships within the larger gene groups.

In general, it remains a remarkable biological observation that the very particular and localized disease phenotype seen in RP can be caused by defects in so many different genes. While studies such as this one can dissect various important differences between the genotype subgroups in RP, the different genetic forms of RP are, overall, similar enough that it supports the theory of a shared underlying pathophysiology (44-46).

### Implicit time

It has long been known that in normal subjects, the retina produces slower electrical responses to dimmer light stimuli.(47) This timing from light onset to response peak is measured as the implicit time, which is recorded in milliseconds. In the context of pathology, a large, healed chorioretinal scar will reduce the amplitude of response without affecting the implicit time, whereas in the case of RP, a decreased amplitude is observed as well as a lengthening of the implicit time.(48) In effect, the chorioretinal scar is a situation where some of the retina is simply missing from an otherwise normally-timed response, while in the case of RP, there is an abnormality of the response timing of the entire retina, as if the entire retina is seeing a dimmer stimulus. More formally, the effect is not simply due to a shortening of cone outer segments and resulting decreased quantal catch of photons, but instead is caused by abnormally low sensitivity of cone phototransduction consistent with a reduction in the amplification of transduction, as well as a slowing of the responses of the inner retina.(49, 50)

There have been hints that a slower (larger) implicit time corresponds to worse disease, for example in sector RP, in which some quadrants of the fundus appear to be normal. When the implicit time is normal or near normal, sector RP has a stationary or more slowly progressive phenotype.

However, patients with very delayed cone flicker implicit time, even if the fundus appears to have “sector RP”, have the progressive form of RP.(51-53) Berson et al took this observation further by noting that a patient’s initial implicit time can numerically help predict the rate of decline of the residual cone response amplitude.(25) Therefore the cone flicker implicit time is a “biomarker” in the sense that is a biologically-derived measurement that has predictive value.

This study validates the use of the cone flicker implicit time as an important biomarker in the rate of progression of RP (Figures 3-4). Longer implicit times are associated with faster rates of ERG amplitude progression, both across the dataset as a whole, and also within the largest genetic subgroups. The relationship may be different in *EYS*, though more data are needed to evaluate this fully. It is interesting that the baseline implicit time is similarly as powerful as the gene name in predicting rate of progression, emphasizing the importance of this biomarker.

Some previous studies have investigated the cone flicker implicit time as an outcome measure, rather than a predictor of disease.(41, 43, 54) One noted that patients with smaller fields may have less delays due to residual foveal cones which are faster.(41) The findings in this study were facilitated by the use of a particular 30 Hz cone flicker ERG protocol (see Methods) that allows recording of responses of lower amplitudes and provides an opportunity to obtain useful amplitude and implicit time data in settings where standard methods may provide more unrecordable signals (e.g. (55)).

### Potential effects of vitamin A and E supplementation in the whole retinitis pigmentosa cohort

It was not our intention to re-evaluate the conclusions of the original study regarding vitamin A and E supplementation in the broader group of all typical RP patients, since the initial goal was to evaluate which genetic subgroups might best respond to vitamin A. However, the data surprisingly indicated that there was no overall robust effect of vitamin A when additional data plus a predictive biomarker were included in the analyses. It is always possible that different data processing or statistical techniques could have uncovered a treatment effect of vitamin A. As described in Methods, we intentionally maintained data processing procedures as close as possible to those used in the original analysis, including use of a minimum starting ERG amplitude when estimating progression rates, and use of an average of both eyes’ amplitudes. We cannot rule out that alternative processing would be more sensitive. An additional floor cutoff at non-baseline visits and explicit modeling of right versus left eye values have been used in later studies, but these techniques were not used in this study in order to most closely replicate the original data processing of the original vitamin A study. As this result was surprising, we invested additional resources into expanding the sample size using subjects in the subsequent trials. These extra subjects did not make a fundamental difference in estimating the vitamin A and E treatment effects but did further enhance the natural history and genetic solutions dataset.

The updated analyses in this study also do not change the structure of the original data. It continues to be the case that the last two years of the original analysis contained less subjects than in the original 4 years, even with additional data from after the original data lock. It has already been noted that the emergence of the vitamin A and E effects occurred in those latter years, where that data was more dispersed (Figure 5 of Berson et al) (9). The lack of a vitamin A treatment effect was seen only when *both* additional data was used from years 5-6, *and* when implicit time was used as a predictive biomarker. The additional 5 and 6 year data brought the data in those years closer to what it had been in years 1-4, in which there were only small differences between the groups. The use of implicit time as a predictor caused the effect to shrink further, because the trial arms were unbalanced with respect to baseline implicit time in a way that coincidentally had made vitamin A look more protective. (The influence of implicit time as a predictive biomarker was not known at the time of the trial.) In summary, there is no robust effect of vitamin A on the progression of RP in this cohort as a whole. The potential negative effect within the *USH2A* subgroup is discussed in the addendum. The modeled negative effect in the *EYS* group may very well be spurious because of the small sample size. The trend toward a potential beneficial effect of vitamin A in the RHO P23H group is intriguing, though only based on 19 subjects and not statistically significant after multiple comparison correction. Most of the additional *RHO* mutations are only represented by only 1 or 2 subjects. Future work could involve binning these mutations by their biochemical properties in order to use a larger fraction of *RHO* subjects to make more robust conclusions.

## Conclusions

Overall, this study demonstrates the systematic differences in severity and progression seen among different genetic subtypes of RP. It further demonstrates how the genetic cause of disease, and the 30Hz ERG implicit time, can have significant predictive power for a patient’s rate of progression. We hope that the lasting contribution of this historical data set will be in helping RP patients and their doctors better understand the severity and expected progression of their disease. Future work may include methods of providing these estimate and predictions in a more accessible format and with refined specificity. Additional endpoints (including visual acuity, visual field, rod-dominant ERG responses) and their progression rates could be evaluated among genetic subgroups as well and may complement data from ongoing prospective natural history studies with additional outcomes such as full-field stimulus threshold testing.(43) Furthermore, the validation of implicit time as a biomarker of disease progression in this large cohort may help with subject selection, subject stratification, and endpoint selection in clinical trials for future experimental therapies (56). Broader adoption of the specialized 30 Hz cone flicker ERG amplitude and implicit time protocol used in this study would likely facilitate the use of the implicit time as a predictive biomarker.

### Subjective clinical recommendations

Patients with RP and their doctors may want to know what is recommended for nutritional supplementation in RP based on this study. When data are complex and do not directly answer all questions of clinical relevance, different investigators who are presented with the same data may draw different conclusions and make different recommendations. Therefore, the rest of this section should be considered opinion rather than direct inference.

In our practice, we have stopped recommending vitamin A supplementation for patients who present with new diagnoses of RP. For patients already on vitamin A, there have been a range of approaches in our practice. One physician’s approach has been to recommend cession immediately or at least by the next visit for all patients, while another physician’s approach has been to allow for continuation under certain circumstances. For example, for patients who have been on vitamin A for many years and feel they are doing well, we have noted a natural tendency for them to want to continue. This seems reasonable as long as yearly liver function tests are performed, but becomes more concerning in the setting of osteopenia or osteoporosis since there is some evidence that high vitamin A intake can worsen bone density (57). A history of renal transplantation can also create additional risk (58). Conversely, we are also concerned that long-time patients who then stop vitamin A supplementation, after having had an experience of slow progression subjectively on vitamin A, may later regret their decision, psychologically, if their disease enters a worse stage. This willingness, by some of our physicians, to allow long-term patients to continue vitamin A supplements when deferring to the patient’s preference, is influenced by our experience that the safety record has been very good (59).

Regarding the potential negative effect of vitamin A supplementation in the subgroup of RP associated with *USH2A* mutations, and to a lesser extent those with *EYS* mutations, it was our impression that the results of the gene-specific subgroup analyses varied widely based on small additions or changes of input data. We speculate that if a large study was conducted in any specific subgroup, then the potential adverse effects would be *un*likely to be replicated. However, notwithstanding the many limitations of any statistical test, the final statistical calculation was well-powered to detect an effect of vitamin A in the *USH2A* subgroup, and the observed effect was adverse. Therefore, for patients with RP associated with *USH2A* mutations who are on vitamin A supplements, we make a recommendation to stop supplementation. The only genetic group for which vitamin A had a trend toward a beneficial effect was for those subjects with the p.Pro23His mutation in *RHO*. The effect size was large, though based on a group of only 19 patients, and was not statistically significant after multiple test correction. While vitamin A supplementation could be considered in this subgroup, there is not high confidence in any recommendation based on this modest amount of data.

For children, retrospective non-randomized data suggests a significant benefit on progression rate in children with typical RP (25); on the other hand, the current study, which was masked and randomized, showed no benefit in adults. These two studies used completely independent data sets on different subjects. We speculate that it is more likely that the retrospective study in children was confounded, compared to the possibilities that there is a different effect in children than adults, or that this study encountered a type II error (failing to identify a true effect). While this speculation is admittedly subjective and not at all certain, we have stopped recommending vitamin A supplementation for children with RP as well.

For vitamin E, we still recommend avoiding high dose supplementation (>30 IU/day). Such supplements are typically marketed as strong anti-antioxidants.

This study does not have any direct impact on the potential effects of DHA or lutein supplementation (5, 10, 11, 54).

In summary, we do not recommend vitamin A or E supplementation for patients with RP. It is possible that further research in subsets of subjects with biochemically distinct mutations would provide additional data. For context, it should be emphasized that other gene-independent, neuroprotective strategies for treatment of patients with IRDs are under study including Rod-derived cone viability factor (RdCVF) (60), N-acetyl cysteine (NAC) (60, 61), and NFE2 like bZIP transcription factor 2 (NFE2L2; previously known as NRF2) (62), which hopefully will provide benefit for RP patients. Gene-specific therapies are of course making great progress as well (63), along with strategies for end stage disease in which there are no rods or cones (e.g. optogenetics) (64).

## Data Availability

Genotype data is available in the Supplementary information, and will be deposited into dbGaP.

## Author contributions

JC: study design, phenotype and genotype data processing and analysis, statistical analyses, writing the manuscript

CWD: study design, phenotype and genotype data processing and analysis, statistical analyses, writing the manuscript

KS: data analysis, writing the manuscript

EP: genotype data processing and variant analysis

MM: phenotype and genotype data processing

EZ: genotype data processing and variant analysis

YZ: statistical analyses

RH: data analysis, writing the manuscript

KB: genotype data processing and variant analysis

EP: study design, phenotype and genotype data analysis, writing the manuscript, obtaining funding

Co-first author order was determined by the authors’ relative contributions to the content, analysis, and writing of the manuscript.

## Acknowledgements

Funding for this study was provided by the Foundation Fighting Blindness and the National Eye Institute RO1 EY012910 (EAP), R01 EY031036 (JC) R01EY026904 (KMB/EAP) and P30EY014104 (MEEI core support). The original vitamin A/E study was funded by the National Eye Institute and the Foundation Fighting Blindness. Center grants from the Foundation Fighting Blindness over the past 4 decades allowed for the creation and maintenance of the resources used in this study, including the DNA biobank and multiple generations of genetic testing infrastructure. The authors would like to thank the research subjects who contributed to the original clinical trials in our department, most of whom traveled across the country every year for extensive testing as trial participants. We would like to thank Eliot Berson, Ted Dryja, Bernard Rosner, their research groups, and all the original investigators and data managers, including Carol Weigel DiFranco and Shyana Harper, who over the last 30+ years who contributed to the collection and maintenance of the large datasets and the large DNA sample repository that were used for this study. We thank Michael Sandberg for his contributions to implicit time as a predictive biomarker and to the software and hardware used for accurate measurement of low amplitude ERG recordings. We thank the clinical staff of the Inherited Disorders Service (formerly, ERG Service) at Mass Eye and Ear. While we know that Dr. Eliot Berson (1937-2017) would be less than pleased with certain findings of this study, we hope that he would be encouraged by the recent progress in the field which he helped create, and by the efforts of the doctors and scientists that he trained who share his commitment toward developing better therapies for patients with inherited retinal diseases.

## Supplemental Methods

### ERG data processing

Data quality flags produced at the time of data acquisition were used to censor data marked as unreliable. A natural log transformation was applied to normalize the distribution of 30 Hz cone flicker ERG amplitudes. Remaining reliable data from right and left eyes were averaged. The amplitudes were intentionally not adjusted for refractive error, to avoid systemically adjusting the averages of certain genotypes which typically have larger refractive errors. Each trial had two baseline visits for each subject, whose values were averaged into a single visit to produce a dataset with only one set of values per year for each subject. Follow-up visit times since the merged baseline visit were rounded to the nearest year. The rate of decline for subjects with very small baseline ERG amplitudes cannot be accurately determined due to higher test-retest variability (“floor effect”). Therefore for the subset of regression analyses that modeled ERG decline over time, a “higher amplitude cohort” was used with baseline amplitudes of at least 0.68 microvolts. For cross-sectional studies, data points from all subjects were used without filtering for a higher starting amplitude. The baseline cone flicker implicit time (alone, and crossed with time) were rescaled to a mean of zero and used as a predictive factor as described. Implicit times from non-baseline visits were not used in the analysis.

### Longitudinal analyses in SAS

A mixed model using SAS version 3.6 was initially used for regression modeling of longitudinal outcomes, to most closely approximate the analysis in the original paper (9). (However, the software used in the SAS statistical analysis package has been upgraded significantly since the early 1990s, and the original mainframe version is no longer available.) A mixed model was used with subject as random variable, and the rounded year as a class variable, “time”. The 2×2 factorial treatment design requires additional coding compared to designs with independent treatment arms; dummy variables for treatment group and treatment group * time were constructed. Treatment groups were: group 1 as A+/E-, group 2 A-/E-(reference), group 3 A+/E+, group 4 A-/E+, where “+” = “high” and “-” = “trace”. (The reference group, which would be the first or last group by convention, was chosen as “group 2” for historical reasons only.) These dummy variables allowed for the use of linear contrasts to model effects of vitamin A and E, as was done for the original study, and as is recommended for 2×2 factorial designs (9, 65). Sample SAS code is given below:

proc mixed data=use;

class year ;

model log30HzERGOU = year_numeric group1 group3 group4 year_times_group1 year_times_group3 year_times_group4 /solution;

random intercept /subject=person_id;

estimate “A effect” year_times_group1 0.5 year_times_group3 0.5 year_times_group4 -0.5 ;

estimate “E effect” year_times_group1 -0.5 year_times_group3 0.5 year_times_group4 0.5 ;

estimate “interaction” year_times_group1 1 year_times_group3 -1 year_times_group4 1 ;

The covariance matrix between study years was inspected and had smoothly decreasing values moving away from the diagonal with range 0.97-0.90, and therefore a random intercept model was used, rather than a repeated measures model with a structured covariance matrix.

For power calculations for longitudinal progression rates within gene subgroups, power was estimated using the sample size formula: 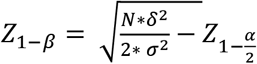, where β is the power corresponding to the z-score of a normal distribution, N is the total sample size, *δ*is the effect size for the yearly rates of change, *σ*^2^is the variance of the estimated slope which equals the sum of between-subject variability and within-subject variability, α is the significance level which is calculated as 5%. The 2 in the denominator reflects the two comparison groups (with and without treatment), for similarly sized groups.

Rates of change are presented in loge units/year of remaining function. This rate represents the speed of the exponential decay. To convert to percent per year, there is an approximation (with <10% error for values less than -0.18 loge units) that, for example, -0.10 loge units/year = -10%/year. The accurate conversion formula is PercentPerYear = 100*(exp(LogsPerYear-1). For example, using this formula, -0.18 loge units per year equals -16.5% per year of remaining function.

### Longitudinal analyses in R

Analyses were performed in version 4.1.3 using the Lmer package. Sample R code is below. “Year” was the study year. “person_id” was the subject id. “Trt” was defined as a factor representing each of the four treatment groups; the reference group was A-/E- and the last three coefficients in the contrast coefficient vector below represent the A+/E-, A+/E+, and A-/E+ groups, respectively.

model = lmer(log30HzERGOU ∼ trt*year + (1|person_id), dataset)

tA = glht(model, linfct = matrix(c(0, 0, 0, 0, 0, 0.5, 0.5, -0.5), 1))

tE = glht(model, linfct = matrix(c(0, 0, 0, 0, 0, -0.5, 0.5, 0.5), 1))

tI = glht(model, linfct = matrix(c(0, 0, 0, 0, 0, 1, -1, 1), 1))

summary(tA)

summary(tE)

summary(tI)

Gene-specific models were also created in R using a similar model. For the model using propensity score matching (Table 2), matching was implemented using the MatchIt package/function in R. For the outlier detection of baseline implicit time described in the text, boxplot.stats()$out was used to detect outliers.

### Homogeneity testing

For homogeneity testing between the three clinical trial data sources, we assessed for homogeneity of the three data sets using the following linear regression model: Ln(ERG amplitude) = b_0_ + b_1_*treatment group + b_2_*year + b_3_*source + b_4_*baseline implicit time + b_5_*treatment group*year + b_6_*source*year + b_7_* baseline implicit time *year, where the b coefficients are fit by the regression model, and “source” is the variable representing one of three clinical trial data sources. The beta described in the text to determine homogeneity is b_6_.

