## Supplemental Table 2 for "Natural history of retinitis pigmentosa based on genotype, vitamin A/E supplementation, and an electroretinogram biomarker"

| Gene | percent per year, this study | percent per year, prior studies |
|---|---|---|
| EYS | -14.7 ± 2.5 | |
| PRPF31 | -9.2 ± 2.0 | -9.2 |
| RHO | -6.6 ± 1.4 | -8.7 |
| RPGR* | -8.6 ± 1.6 | -7.1 |
| USH2A | -13.8 ± 1.3 | -13.2 |

Supplemental Table 2. Progression rate of the 30 Hz cone flicker ERG amplitude of the largest gene groups, in percent per year of remaining function, ± 95% confidence interval of the mean. *The *RPGR* cohort was restricted to an RP phenotype in this cohort, but not in the prior study.
